# Early empirical assessment of the N501Y mutant strains of SARS-CoV-2 in the United Kingdom, October to November 2020

**DOI:** 10.1101/2020.12.20.20248581

**Authors:** Kathy Leung, Marcus HH Shum, Gabriel M Leung, Tommy TY Lam, Joseph T Wu

## Abstract

Two new SARS-CoV-2 lineages with the N501Y mutation in the receptor binding domain of the spike protein have rapidly become prevalent in the UK. We estimated that the earlier 501Y lineage without amino acid deletion Δ69/Δ70 circulating mainly between early September to mid-November was 10% (6-13%) more transmissible than the 501N lineage, and the currently dominant 501Y lineage with amino acid deletion Δ69/Δ70 circulating since late September was 75% (70-80%) more transmissible than the 501N lineage.

## Main text

Two new SARS-CoV-2 lineages carrying the amino acid substitution N501Y in the receptor binding domain (RBD) of the spike protein (S protein) have rapidly become prevalent in the UK. The earlier 501Y lineage (501Y Variant 1) cocirculated with the 501N lineage between early September and mid-November in Wales, where the prevalence never exceeded 2%. However, a later 501Y lineage (501Y Variant 2) started to cocirculate with the 501N lineage in England since late September and has since become the dominant lineage from late November. In the UK, the prevalence of the latter 501Y Variant 2 lineage has increased from 0.1% in early October to 49.7% in the late November. Of note, the N501Y mutation co-occurs with several mutations in the ORF1a, ORF8, N and the S protein in 501Y Variant 2, including two amino acid deletions Δ69 and Δ70.

The rapid spread of 501Y Variant 2 suggests it may have a transmission advantage over the 501N lineage. Structural biological studies of the SARS-CoV-2 RBD offer insights proposing that 501Y may increase ACE2 binding [1] and that the open state conformation of the 501Y S protein [2] is associated with more efficient viral entry and infection. Epidemiologically however, there has been limited assessment to date investigating whether any of these mutations may have changed transmissibility [3]. Here we adopted our previous epidemiological framework for relative fitness inference of co-circulating pathogen strains, which has been applied on influenza viruses [4] and SARS-CoV-2 614D/G strains [5], to characterize the comparative transmissibility of the 501Y Variant 1 and Variant 2 strains.

### Reconstructing the phylogeny of 501N, 501Y Variant 1 and 501Y Variant 2 in the UK

We downloaded the multiple sequence alignment of complete (and nearly complete) genomes of SARS-CoV-2 from the GISAID database (www.gisaid.org) on 14 December 2020. We extracted all those viral genomes carrying 501Y in the translated spike protein sequences, and analyzed them with other closely related virus strains (identified through BLAST search) in the global phylogeny. The resultant phylogeny built with the maximum likelihood method and GTR substitution model using FastTree version 2.1 [6] is shown in Figure 1. It indicates that the recent 501Y strains in UK since August/September 2020 forming two lineages emerged from the 20B clade (NextStrain nomenclature). Both lineages have clear geographical separation in Wales vs England. The first 501Y lineage (501Y Variant 1) appeared in Wales in early September and persisted through November. The second 501Y lineage (501Y Variant 2) appeared in England in late September and largely expanded to become the predominant lineage in the region since late November. Notably, the 501Y Variant 2 lineage has carried a number of common amino acid mutations including the deletion of two amino acid at 69th and 70th residues (Δ69/Δ70) and the deletion of single amino acid at 144th residue on the spike protein. Globally, two other lineages with 501Y (without Δ69/Δ70) have been detected in Australia and South Africa circulating during June-July and October-November 2020, respectively.

**Figure 1.**
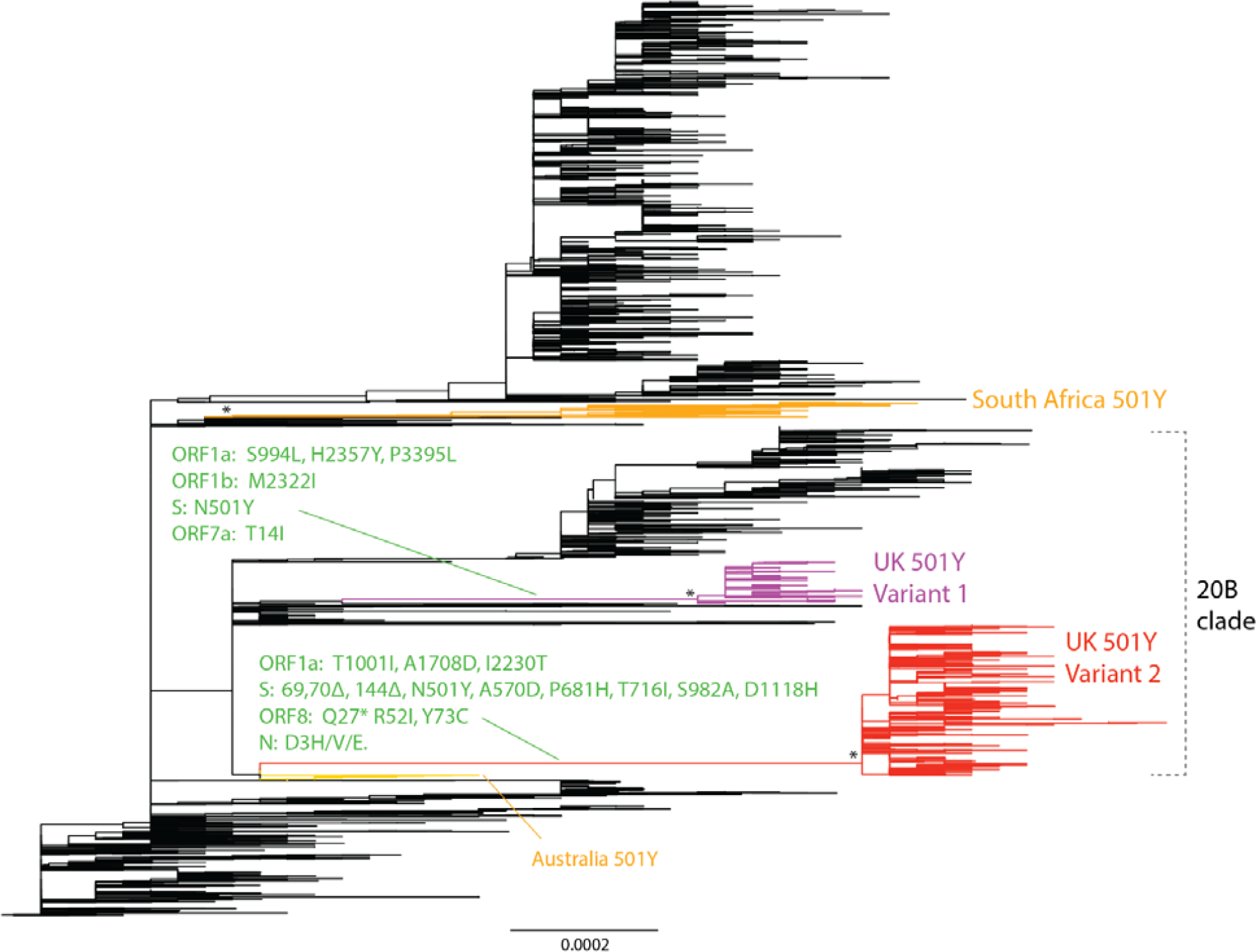
Phylogeny of SARS-CoV-2 showing the emergence of 501Y lineages in UK and other regions. The maximum likelihood tree was built from 7,003 genome nucleotide sequences of SARS-CoV-2. The UK 501Y Variant 1 and 2 lineages are coloured in purple and red respectively. The South African and Australian 501Y lineages are in orange colour. Mutations at the preceding branches of UK 501Y Variant 1 and 2 are indicated in green. Some 501Y strains in sporadic emergence (including those in Spain, USA etc) without establishment of lineage of more than ten sequences are not shown. The asterisks refer to the >0.98 topology supports for the monophyletic grouping of the 501Y lineages. Branch scale is shown at the bottom of the tree in the unit of substitutions per site.

To include more sequences for the study of virus fitness, we extended our search of more 501Y Variant 1 and Variant 2 sequences in the latest GISAID dataset downloaded on 19 December 2020, including both the complete genomes and the partial ones covering spike genes.

### Comparative transmissibility of 501Y Variant 1 and Variant 2

We applied a fitness inference framework to the sequence data collected from the UK between September 29 and November 16 during the cocirculation period of the three strains (see Supplementary Information for details). We assumed that the N501Y mutation and Δ69/Δ70 deletions characterize the three strains 501N, 501Y Variant 1 and 501Y Variant 2, but their differential transmissibility (if any) might be attributable to the combination of N501Y and other mutations including Δ69/Δ70 deletions acquired in the emergence of 501Y Variant 1 and 2 lineages (Figure 1). For conciseness, we used *N, Y*1 and *Y*2 to denote the three strains. We defined the comparative transmissibility of any two strains as the ratio of their basic reproductive numbers. That is, the comparative transmissibility of strains *Y*1 and *Y*2 with respective to strain *N* was 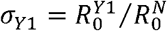 and 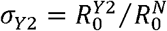, respectively.

Using confirmed deaths (adjusted for the delay between onset and death [7]) as the proxy for the COVID-19 epidemic curve [8], we estimated that *σ*_*Y*1_ was 1.10 (95% CrI 1.06-1.13) and *σ*_*Y*2_ was 1.75 (1.70-1.80). That is, the basic reproductive number of the 501Y Variant 1 and Variant 2 was 10% (6-13%) and 75% (70-80%) higher than that of the 501N stain, respectively.

The fitted model was largely congruent with the observed proportions of the three strains over time, except in the last week of November 3-10 during which 501Y Variant 1 cocirculated with 501N and 501Y Variant 2 (Figure 2). Since 501Y Variant 1 mainly cocirculated with 501N in Wales, we performed a sensitivity analysis on sequence data from Wales only. We estimated *σ*_*Y*1_was 1.14 (95% CrI 1.11-1.19) but were not able to estimate *σ*_*Y*2_ because currently there are only two 501Y Variant 2 sequences sampled before November 30 from Wales from our dataset.

**Figure 2.**
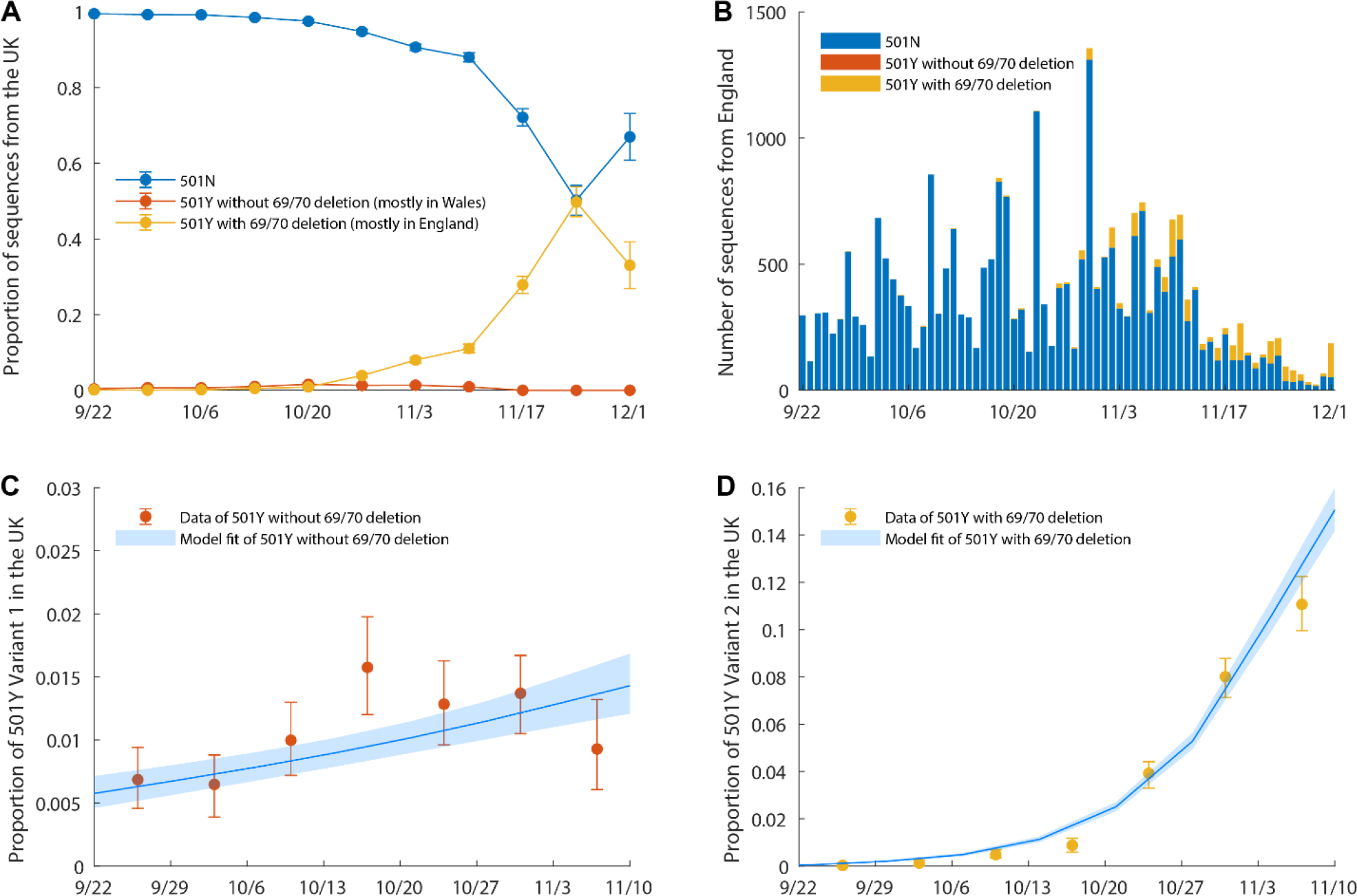
The weekly proportion of 501N, 501Y Variant 1 and 501Y Variant 2 sequences between late September and early November when three strains cocirculated. The time series of confirmed COVID-19 deaths in the UK was used in the estimation. (A) The proportion of sequences submitted by the UK between September 22 and December 1. (B) The number of sequences submitted by England between September 22 and December 1. (C) The proportion of 501Y Variant 1 sequences during the cocirculation period between September 22 and November 16. (D) The proportion of 501Y Variant 2 sequences. The circles and error bars indicated the observed proportion with 95% multinomial CIs among sequence data. The blue lines and shades indicated the posterior mean and 95% CrI of the estimates.

## Discussion

Our findings indicate that the 501Y Variant 2 is 75% more transmissible than the 501N strain in the UK; thus the *R*_0_ would be 1.75 times that of the 501N strain. This variant has also become the dominant strain in England. These observations would imply more rapid and stringent control measures would be necessary to suppress spread, which is precisely what the UK government effected on December 19, including the addition of a new tier 4 set of restrictions [9].

Outside the UK, only a small number of 501Y Variant 2 has been identified (e.g., in Denmark), but it is unclear whether they were exports from the UK until more sequence data become available. Although sporadic spread of the 501Y mutation occurred in Wales and elsewhere (e.g., Australia, Spain, and US), not all variants with 501Y have become prominent. In South Africa, a new variant with 501Y but not Δ69/Δ70 has emerged and spread rapidly since late October [10]. Our phylogenetic analyses show that the South African variant is genetically distant from 501Y Variant 2 and has many mutations not shared with 501Y Variant 2, suggesting the role of 501Y is more complex. Other mutations (such as Δ69/Δ70 and D1118H of the S protein) may account for the increased transmissibility of 501Y Variant 2. Future studies of their individual and combinatorial effects to the viral phenotypes are warranted.

Further work should clarify the role, if any, of increased mobility and population mixing that was concurrent with the circulation of the 501Y Variant 2 in explaining higher transmissibility. Assessment of clinical severity changes associated with these variants would require several more weeks of close and careful observation. Finally, given the numerous mutations associated with 501Y variants, thus the potential for antigenic changes, immuno-genomic surveillance should be stepped up to identify instances of reinfection in previous confirmed patients as well as breakthrough infections amongst vaccines.

## Data sharing statement

We collated all data from publicly available data sources. All data included in the analyses are available in the main text or the supplementary materials.

## Acknowledgement

We thank colleagues who have shared the SARS-CoV-2 sequences in GISAID (www.gisaid.org).

## Contributors

KL, TTYL, JTW and GML designed the experiments. KL, MHHS and TTYL collected data. TTYL and MHHS performed sequence alignment and phylogenetic analysis. KL and JTW analyzed epidemiological data. KL, JTW, TTYL, and GML interpreted the results and wrote the manuscript.

## Supplementary Information

### Inference framework to estimate comparative transmissibility

We assumed that the N501Y mutation and Δ69/Δ70 deletions characterize the three strains 501N, 501Y Variant 1 and 501Y Variant 2, but their differential transmissibility (if any) might be attributable to the combination of N501Y and other mutations including Δ69/Δ70 deletions acquired in the emergence of 501Y Variant 1 and 2 lineages (Figure 1). For conciseness, we used *N, Y*1 and *Y*2 to denote the three strains. We defined the comparative transmissibility of any two strains as the ratio of their basic reproductive numbers. That is, the comparative transmissibility of strains *Y*1 and *Y*2 with respective to strain *N* was 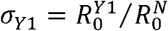 and 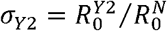, respectively.

We formulated a framework to infer *σ*_*Y*1_ and *σ*_*Y*2_ under the following base case assumptions: (1) the three strains co-circulated locally during our study period (September 22 to November 16, 2020); (2) non-pharmaceutical interventions (NPIs) had the same effect on all three strains; (3) the probability that an infected person was selected for viral sequencing did not depend on which strain he/she was infected with; (4) recovery from infection with any strain provided protection against reinfection of all strains during our study period; (5) age-specific susceptibility to infection (if any) was the same for all three strains; and (6) after community transmission of strains *Y*1 and *Y*2 have been established, the effect of further de novo emergence on their prevalence was negligible.

Under these base case assumptions [4, 5], the next generation matrix (NGM) of infections by strains *Y*1 and *Y*2 were *σ*_*Y*1_ and *σ*_*Y*2_ times that of the strain *N*. Let *ρ*_*j*_(*t*) be the proportion of strain *j* among all new COVID-19 infections generated at time. As the pandemic unfolds, *ρ*_*j*_(*t*) would increase monotonically towards 1 if strain *j* had the highest comparative transmissibility. In our previous work [4, 5], we have shown that *ρ*_*j*_(*t*) can be well-approximated using the equation:

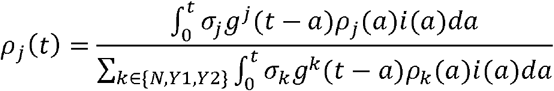

Where *i*(*t*) was the total incidence rate (i.e., including three strains),*g*^*j*^ was the generation time distribution for strain *j*. In the base case, we assumed that all three strains had the same generation time distribution with mean 5.4 days [7]. Let 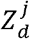 be the number of SARS-CoV-2 sequences sampled on day *d* that was strain *j*, and *ĩ*(*t*) be a reliable proxy of the incidence rate *i*(*t*). We substitute *i*(*t*) with *ĩ*(*t*)and denote the resulting approximation of *ρ*_*j*_(*t*) by 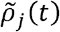. The comparative transmissibility of strains *Y*1 and *Y*2 with respect to strain *N* (i.e., *σ*_*Y*1_ and *σ*_*Y*2_) were estimated using the following likelihood function:

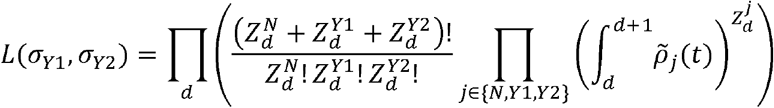

The statistical inference was performed in a Bayesian framework with non-informative (flat) priors using Markov Chain Monte Carlo.

